# Sustainable and portable CRISPR-based diagnostics for high-sensitivity Mpox detection

**DOI:** 10.1101/2024.11.20.24317678

**Authors:** Rika Hirano, Kazuto Yoshimi, Koji Asano, Kohei Takeshita, Ken J. Ishii, Kei Sato, Tomoji Mashimo

## Abstract

Mpox has emerged as a critical public health challenge, creating an urgent need for rapid, reliable, and field-deployable diagnostic tools for outbreak settings. Here, we present Kairo-CONAN, a novel CRISPR-Cas3-based point-of-care (POC) diagnostic platform for Mpox, engineered for sustainability and portability. This system leverages a disposable hand warmer (Kairo) as a stable heat source and incorporates freeze-dried reagents for ambient temperature stability, enabling device-free, sensitive detection through lateral flow assay strips. Utilizing CRISPR-Cas3’s unique DNA-targeting and cleavage properties, we optimized probe DNA configurations for high specificity and designed clade-specific target crRNAs. Kairo-CONAN demonstrated rapid, high-sensitivity, and specific detection of Mpox virus (MPXV) DNA across multiple clades, including Clade Ia (Congo), Clade Ib (synthetic DNA), and Clade IIb (Tokyo). By addressing logistical and environmental challenges, Kairo-CONAN offers a sustainable, cost-effective, and field-adapted solution for infectious disease diagnostics, aligning with the 100-day mission framework to enhance global outbreak response efforts.

## Introduction

The recent global resurgence of monkeypox (Mpox), caused by the orthopoxvirus Monkeypox virus (MPXV), has revealed critical public health challenges and underscored the urgent need for rapid, reliable, and accessible diagnostic tools, particularly in outbreak-prone regions^1–4^. Traditionally endemic to Central and West Africa, Mpox has spread significantly since 2022, with cases reported across multiple continents^5,6^. In response, the World Health Organization (WHO) classified Mpox as a Public Health Emergency of International Concern (PHEIC), emphasizing the need for field-deployable diagnostics to effectively monitor and contain outbreaks^7^. Addressing this, the G7 initiated the 100 Days Mission (100DM), calling for the development of portable, accurate, and device-free diagnostic tools to enable testing in resource-limited settings (https://ippsecretariat.org/). Current diagnostic methods for Mpox face substantial limitations. PCR-based testing, while highly sensitive and specific, requires specialized laboratory infrastructure, trained personnel, and a stable power supply—resources often unavailable in remote or emergency settings^8,9^. Conversely, rapid antigen tests, though portable, lack the sensitivity and specificity required for accurate detection in diverse field environments^10,11^. These limitations highlight an urgent need for innovative diagnostic platforms that combine high sensitivity, specificity, portability, and ease of use for low-resource settings.

CRISPR-based diagnostics have emerged as a promising solution, offering high sensitivity and specificity with rapid, point-of-care potential without reliance on complex equipment^12–15^. While CRISPR systems such as Cas12a and Cas13 are widely utilized, CRISPR-Cas3 offers unique advantages, including the ability to target longer DNA sequences and perform collateral cleavage, enhancing specificity and reducing background noise^17^. These properties are particularly beneficial for precise viral detection. Our previous work with the CRISPR-Cas3-based CONAN platform^16^ demonstrated high specificity for nucleic acid detection, inspiring its adaptation for field-based Mpox diagnostics.

In this study, we present Kairo-CONAN, a CRISPR-Cas3-based diagnostic system optimized specifically for Mpox detection. Designed for sustainability and portability, Kairo-CONAN utilizes a disposable hand warmer ("Kairo" in Japan) as a stable, device-free heat source^17^ and incorporates freeze-dried reagents for room-temperature stability^18^. To maximize detection accuracy, we developed clade-specific crRNAs and optimized probe DNA sequences, enabling targeted detection across Mpox clades Ia, Ib, and IIb^19^. Validation of Kairo-CONAN demonstrated robust performance across these clades, highlighting its adaptability and reliability for pandemic preparedness. By combining the specificity of CRISPR-Cas3 with a sustainable, device-free design, Kairo-CONAN offers a novel approach to infectious disease diagnostics. This platform aligns with the G7’s 100DM framework by addressing logistical and environmental barriers, supporting rapid deployment in outbreak settings, and strengthening global diagnostic preparedness for Mpox and other emerging infectious diseases.

## Results

### CRISPR-Cas3 mediated diagnostics for monkeypox virus (MPXV)

We optimized the production of functional Cascade complexes by individually purifying Cas components (Cas5, Cas6, Cas7, Cas8, and Cas11) using a baculovirus expression system in *Spodoptera frugiperda* (Sf9) insect cells^20^. The purified components were then assembled with synthetic crRNA *in vitro* to form *in vitro* assembled Cascade complexes (ivCascade) ^21^. This streamlined approach eliminates the need for recombinant Cascade complex (rnCascade) purification from *Escherichia coli* as previously reported^16^, significantly simplifying CRISPR-Cas3 diagnostic workflows. The ivCascade production method enables rapid nucleic acid detection, which is particularly advantageous during pandemic scenarios such as COVID-19 and Mpox.

To validate the CRISPR-Cas3-based CONAN platform for MPXV detection, target sequences were designed in accordance with CDC diagnostic guidelines^9^ (Fig. 1A). Using the targeting crRNAs, CRISPR-Cas3 collateral cleavage activity specific to MPXV DNA was observed after 10 minutes of incubation at 37°C (Fig. 1B). Coupling this assay with recombinase polymerase amplification (RPA) enabled detection of MPXV sequences at concentrations as low as 100 aM (approximately dozens of copies), demonstrating the assay’s sensitivity and utility for MPXV detection (Fig. 1C).

**Figure 1.**
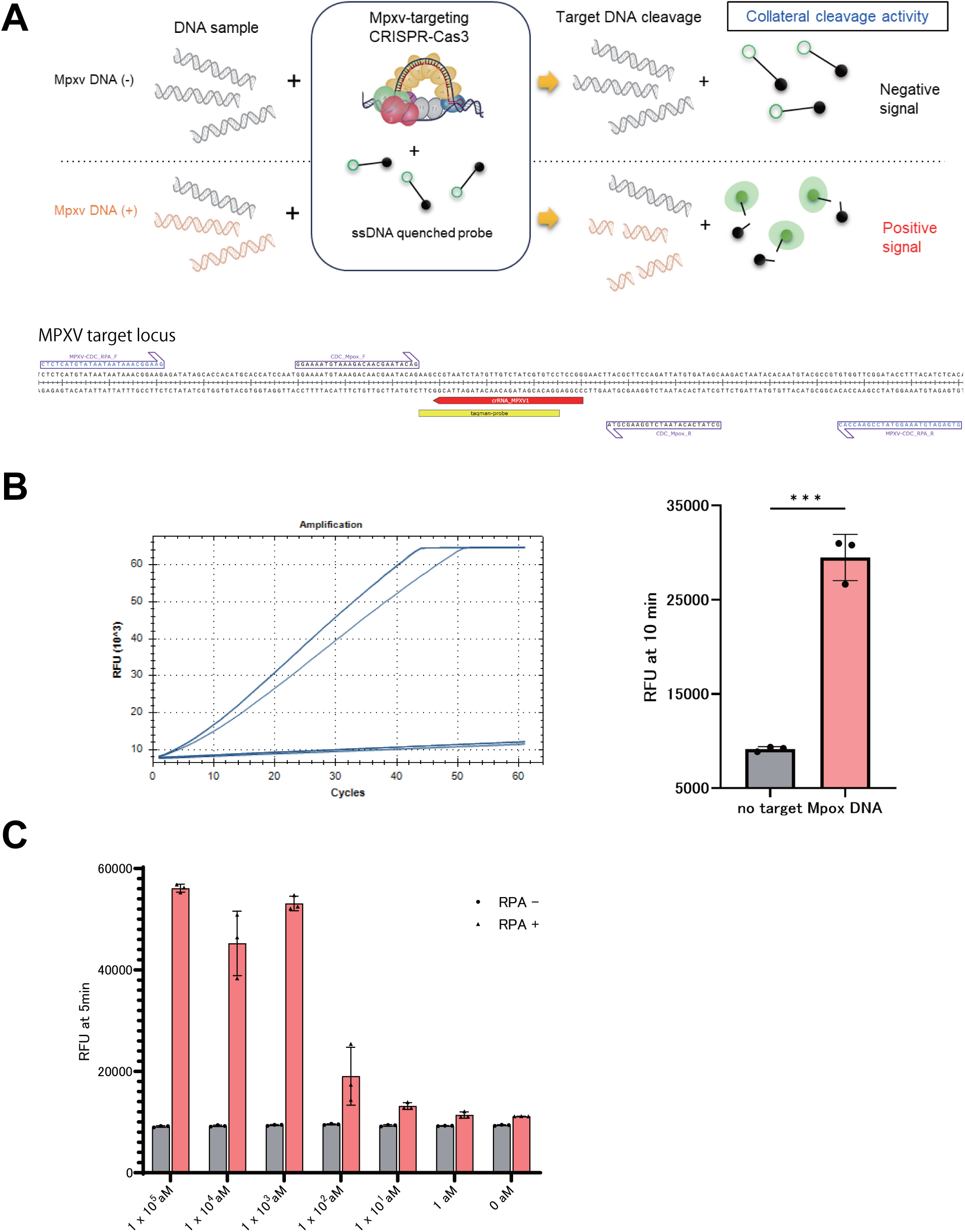
Detection of Monkeypox Virus by CIRPSR-Cas3 based diagnistics. A) Schematic representation of CRISPR-Cas3-based CONAN platform for MPXV DNA detection using in vitro assembled Cascade complexes (ivCascade) and Cas3 protein. A crRNA, common #1, was designed to target the same genomic region as the CDC RT-qPCR primer set. B) Collateral ssDNA cleavage activity measured sequentially by incubation of EcoCas3-EcoCascade/crRNA complex with a 100-bp dsDNA activator containing a target sequence in the MPXV genome at 37 °C. CRISPR-Cas3 mediated collateral ssDNA cleavage after targeting *MPXV*-dsDNA in fragments, quantitatively represented by relative fluorescent units (RFU) after 10 minutes (right graph). Means (n = 3), and standard deviations. Statistical analysis was performed using the two-tailed Student’s t-test. C) The CONAN assay with isothermal RPA amplicon products (red) detected several copies of the *MPXV* activator fragments (10 aM); RFU at 10 min. Means (n = 3), and standard deviations.

### Device-free nucleic acid detection using disposable hand warmers (Kairo)

During Mpox and other pandemics, the lack of approved antigen tests underscores the need for accurate diagnostics in outbreak regions with limited infrastructure. To address this, we developed a device-free nucleic acid detection system using a disposable hand warmer (Kairo) composed of activated carbon as a heat source (Fig. 2A). Various models were tested (Supplementary Fig. 1A and 1B), and a specific Kairo model stabilized at approximately 37°C within 10 minutes (Supplementary Fig. 1A), enabling effective nucleic acid amplification. Using a preheated Kairo, RPA reactions achieved comparable performance to a 37°C incubator, with detectable signals within 10 minutes of the target sequence (Fig. 2B). Collateral cleavage activity of CRISPR-Cas3 targeting the EMX1 sequence was similarly validated with Kairo, showing equivalent activity to standard incubation conditions (Fig. 2C). Lateral flow detection further demonstrated the successful integration of Kairo into a fully device-free workflow (Fig. 2C).

**Figure 2.**
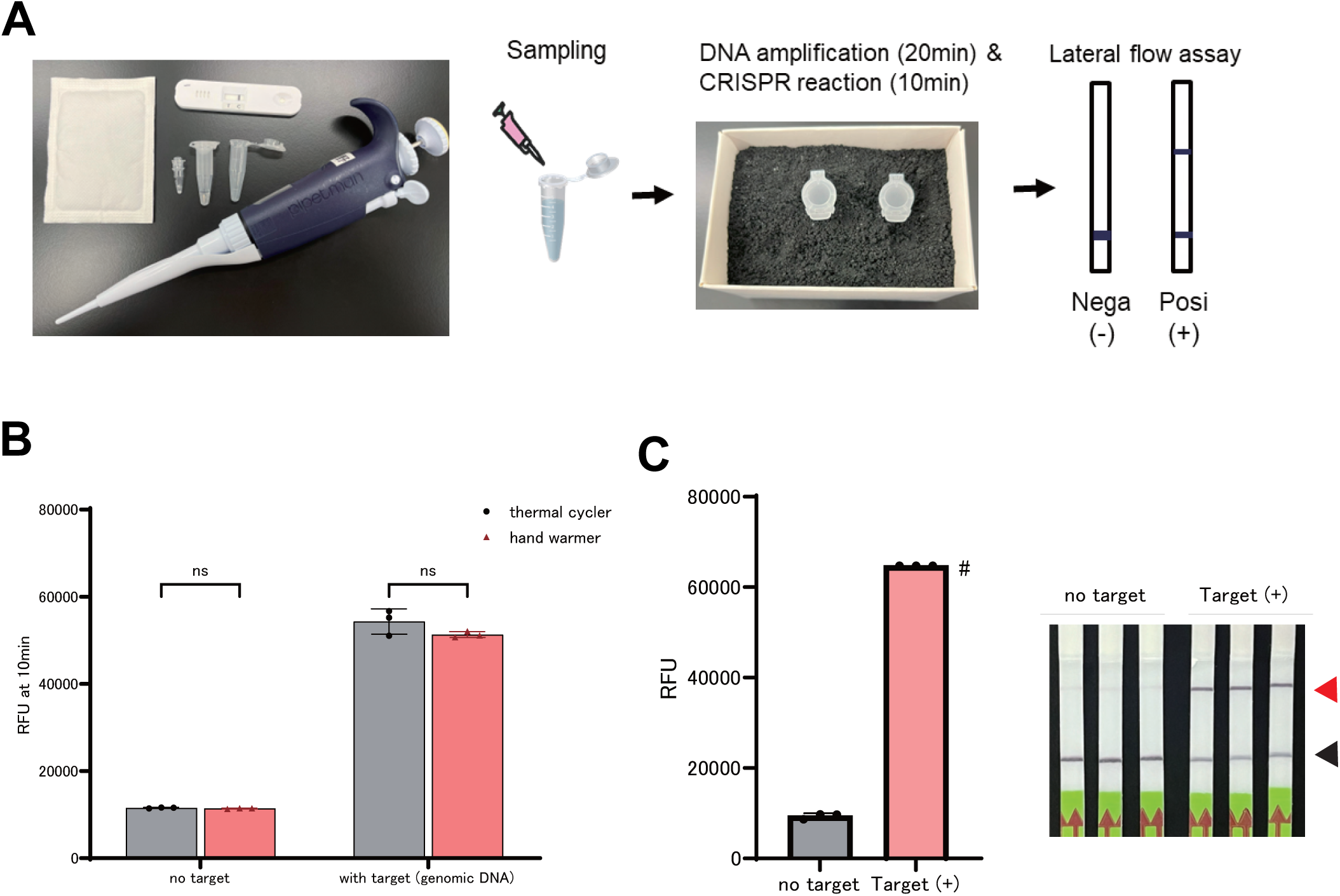
Device-Free CONAN Using Disposable Hand Warmers (Kairo) A) Setup diagram illustrating the use of hand warmer (Kairo), as an alternative and disposable heat source for RPA and CONAN assay. B) Collateral cleavage activity of EMX1-target CRISPR-Cas3 after the RPA amplification with thermal-cycler or hand warmer incubation. ns: no significance. C) Collateral cleavage activity and lateral flow detection of CRISPR-Cas3 after the CONAN reaction with hand warmer incubation for 10 min. #: Saturated signals on CFX Connect device. Positive (red arrow) and negative (black arrow) bands for CONAN

We confirmed the applicability of Kairo for RT-RPA, enabling detection of RNA viruses such as SARS-CoV-2 (Supplementary Fig. 2A), and extended the system to CRISPR-Cas12a diagnostics with consistent results (Supplementary Fig. 2B). Notably, Kairo-based reactions performed reliably even at 4°C, demonstrating the feasibility of nucleic acid detection in diverse environments, including cold and off-grid settings (Supplementary Fig. 3).

### Lyophilized CRISPR-Cas3 components for field adaptation

To develop a device-free CRISPR diagnostic suitable for field use, we optimized freeze-drying conditions for CRISPR-Cas3 components to enable stable storage at room temperature ^18^ (Fig. 3A). Initially, we examined the effect of trehalose, a common cryoprotectant, on the activity of Cas3 and EMX1-targeted rnCascade ^16^. When Cas3 or rnCascade was lyophilized with buffers containing different concentrations of trehalose, the buffer containing 10% trehalose maintained the highest CONAN activity (Supplementary Fig. 4A and 4B). This buffer was then applied to ivCascade complexes targeting EMX1 ^21^, and was lyophilized. These lyophilized complexes retained activity comparable to freshly prepared counterparts after freeze-drying and thawing, confirming the stability of Cascade proteins under these conditions (Fig. 3B). CONAN activity was also observed with lyophilized ivCascade complexes targeting the common region of MPXV (Supplementary Fig. 5).

**Figure 3.**
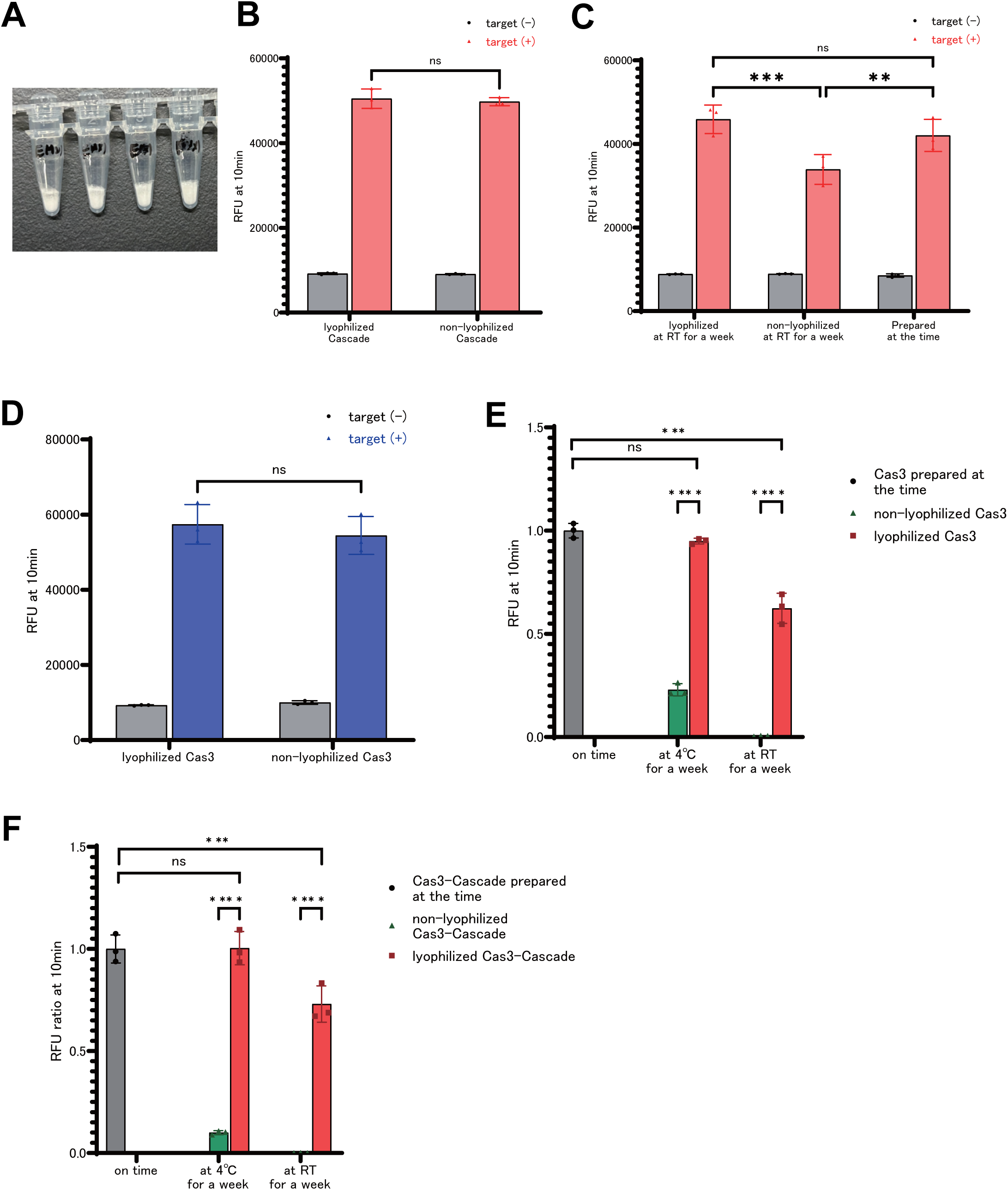
Lyophilization of CRISPR-Cas3 Components for stable storage at room temperature. A) Representative image of lyophilized Cascade protein for EMX1-target with 10% trehalose. B) Collateral cleavage activity of CRISPR-Cas3 with the ivCascade targeting EMX1 before and after lyophilization. ns: no significance. C) Collateral cleavage activity of Cascade stored at room temperature for one week in lyophilized or non-lyophilized. **: p<0.01 ***:p<0.001 D) Collateral cleavage activity of of CRISPR-Cas3 with the Cas3 protein before and after lyophilization. ns: no significance. E) Collateral cleavage activity of CRISPR-Cas3 with lyophilized or non-lyophilized Cas3 stored at 4°C or room temperature for one week. *: p<0.05 ****:p<0.0001 F) Collateral cleavage activity of lyophilized or non-lyophilized Cas3 and Cascade protein mix stored at 4°C or room temperature for one week. *: p<0.05 ****:p<0.0001

We further assessed the stability of lyophilized ivCascade complexes stored at room temperature and 4°C for one week, comparing their activity to non-lyophilized counterparts. Lyophilized complexes exhibited significant cleavage activity under both conditions, often surpassing that of non-lyophilized samples (Fig. 3C). Similarly, Cas3 protein, known for its instability at room temperature and even at 4°C ^22^, was evaluated under lyophilized conditions. When freeze-dried in a 10% trehalose buffer, Cas3 retained complete activity with no detectable loss compared to pre-lyophilization levels (Fig. 3D). After one week of storage, lyophilized Cas3 maintained over 95% of its initial activity at 4°C, whereas its activity in buffer solutions stored at the same temperature dropped by more than 70%. At room temperature, lyophilized Cas3 retained approximately 60% of its activity, while Cas3 in buffer lost all activity (Fig. 3E).

To further enhance the field applicability, we prepared a lyophilized mixture of Cas3-cascade complexes for single-tube functionality. Stability tests showed similar profiles to those observed with Cas3 protein alone: samples stored at 4°C maintained activity levels comparable to freshly prepared solutions, while room temperature storage retained approximately 70% of initial activity (Fig. 3F). These results demonstrate that the lyophilized Cas3-cascade complex is stable under various storage conditions, can be reactivated with water, and is immediately usable, making it suitable for field deployment.

### Optimization of the DNA probes for enhanced sensitivity

While the lyophilized, device-free Kairo-CONAN system enables portable applications, its signal sensitivity may be affected by environmental factors and storage conditions. To enhance CRISPR-Cas3 collateral cleavage activity, we optimized the design of DNA probes used for ssDNA detection. Since Cas3 cleaves single-stranded DNA (ssDNA) with minimal impact on single-stranded RNA (ssRNA) ^22^ we designed RNA probes with two DNA base substitutions to investigate cleavage efficiency. Probes with pyrimidine bases (thymine and cytosine) at the 5’ end demonstrated higher cleavage efficiency compared to probes with purine bases (adenine or guanine), with guanine significantly reducing cleavage activity (Fig. 4A). Heatmap analysis of cleavage activity revealed that although pyrimidine bases at the 5’ end increased cleavage activity, they also induced non-specific cleavage in the absence of target sequences. After screening for optimal signal-to-noise (SN) ratios, sequences such as CC, CA, and TC were identified as having the highest SN ratios (Fig. 4B).

**Figure 4:**
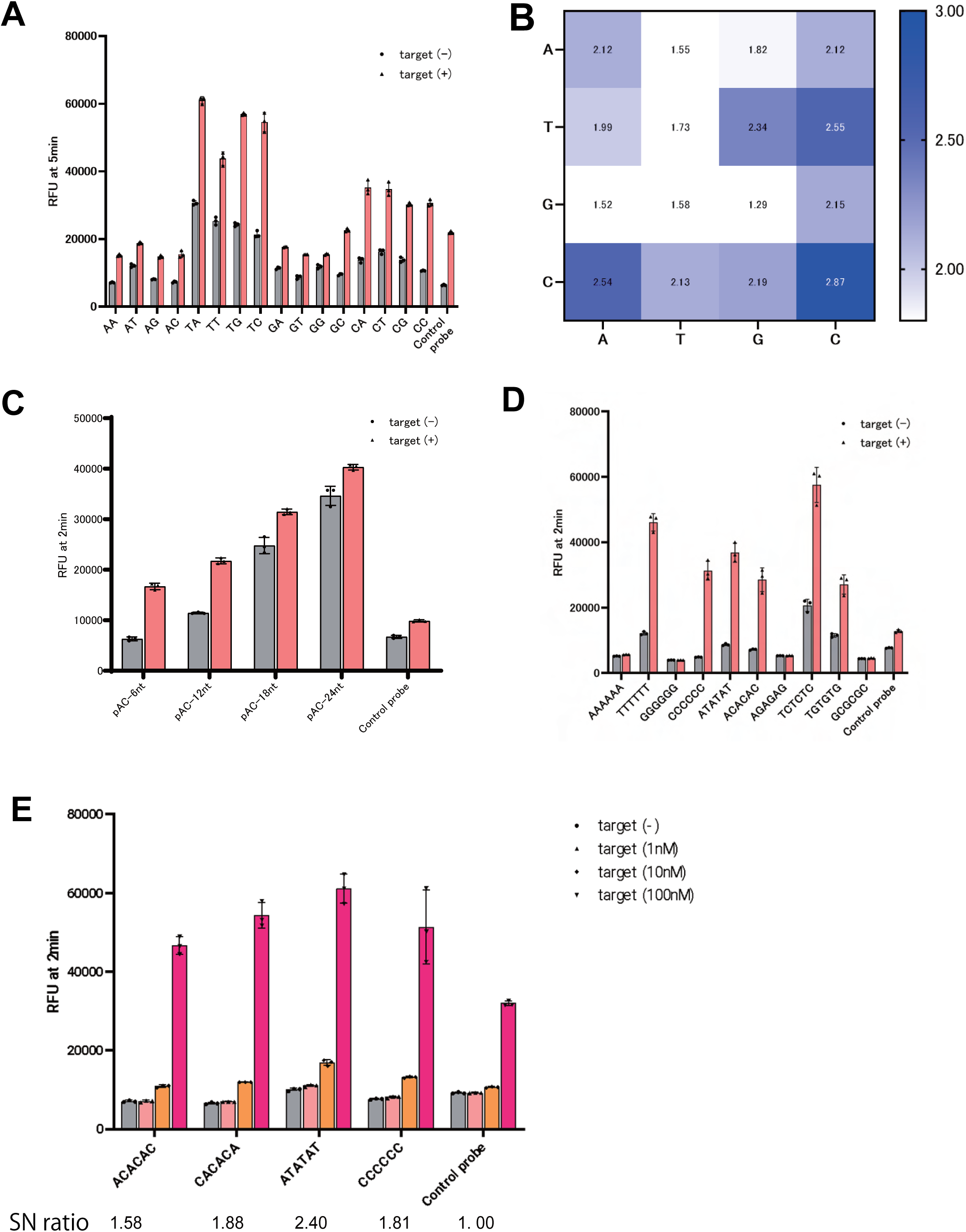
Cas3 ssDNA cleavage preferences for optimizing probes for CONAN. A) Collateral cleavage activity of EMX1 target sequences using the RNA-DNA hybrid probes composed of two deoxynucleotides and the heat map of the collateral cleavage activity of the two base combinations. B) The signal-to-noise ratio in the presence of non-specific cleavage activity without the target. C) Evaluation of the collateral cleavage activity with the different length of adenine and cytosine repeat sequences. D) Assessment of the optimal probe sequence with high signal-to-noise ratio by evaluating collateral cleavage activity of 6-base ssDNA probes with various combinations of four deoxyribonucleotides. E) Evaluation of the collateral cleavage activity for MPXV target with several optimized probes.

To determine the optimal probe length, we tested probes featuring AC sequences repeated at 6-base intervals. While longer probes provided higher signal intensities, they also increased non-specific cleavage, indicating that shorter probes with 6-base sequences were more suitable for improving specificity (Fig. 4C). Further evaluation of probe sequences revealed that probes with six consecutive bases of single or dual pyrimidine bases exhibited high specificity. However, thymine-based probes showed lower SN ratios due to non-specific cleavage, whereas pyrimidine and adenine combinations maintained high activity with reduced non-specific cleavage, achieving improved SN ratios (Fig. 4D).

Through this optimization process, the ideal probe sequence was determined to consist of six bases, combining pyrimidine and adenine bases for enhanced cleavage specificity and SN ratio (Fig. 4E). Sensitivity tests with serial dilutions demonstrated that the optimized probe increased sensitivity by approximately 2.4-fold compared to the GAPDH probe. These optimizations are expected to enhance the overall performance of Kairo-CONAN systems, including Mpox detection, by enabling the detection of lower nucleic acid concentrations and reducing the required RPA incubation times.

### Kairo-CONAN provides diagnostic methods for Mpox pandemic response

Following the COVID-19 pandemic, Mpox re-emerged in Central Africa, particularly in the Democratic Republic of Congo, prompting the WHO to classify it as a Public Health Emergency of International Concern (PHEIC) in August 2024^7^. In response, the G7’s 100 Days Mission (100DM) initiative was launched to accelerate the deployment of diagnostics, vaccines, and treatments within 100 days of a PHEIC declaration, aiming to ensure rapid and effective medical countermeasures (https://ippsecretariat.org/). Kairo-CONAN, a lyophilized, device-free CRISPR-Cas3 diagnostic platform, demonstrates potential as a robust solution for low-resource settings during pandemics. To evaluate its adaptability, we validated Clade-specific detection using newly released Clade Ib sequences^23,24^. Sequences for Mpox Clades Ia, Ib, and IIb were sourced from the CDC and NCBI databases, and target sequences were designed and synthesized as crRNAs^8,9,23,24^ (Fig. 5A). The synthesis process required approximately two weeks, demonstrating the platform’s readiness for rapid deployment after their sequence release.

**Figure 5:**
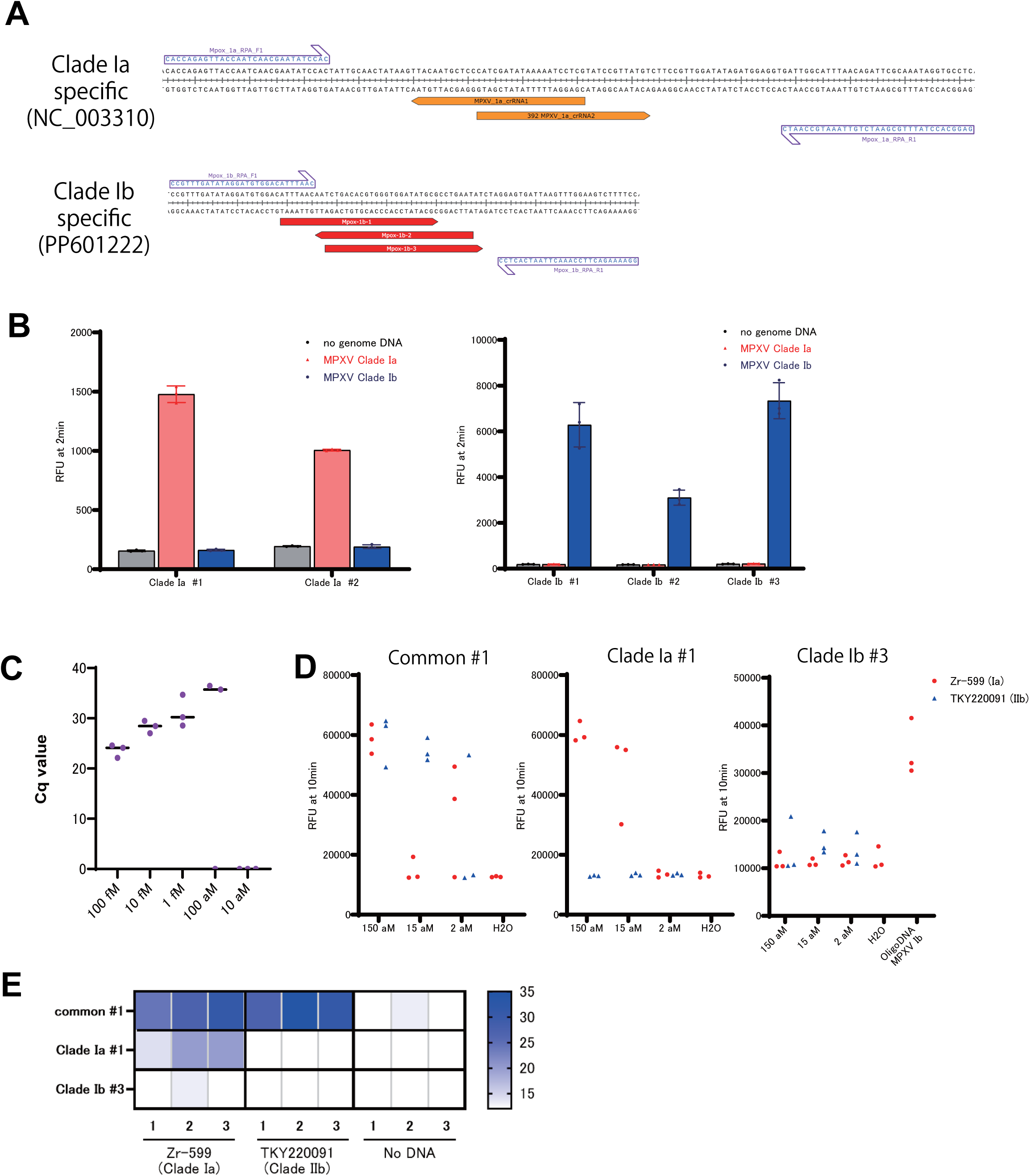
Clade-Specific Detection of MPXV. A) Design of primers for RPA and targeted crRNA candidates for MPXV Clade Ia and Clade Ib. B) Target-specific collateral cleavage activity of MPXV-target CRISPR-Cas3 to the MPXV Clade Ia and Ib amplicons by RPA reaction. C) The limit of detection (LoD) of the US CDC’s RT-qPCR assay amplification of MPXV. Cq, cycle quantification value. D) LoD of Kairo-CONAN-based assays for CDC common, Clade Ia and Ib specific crRNAs to the genomic DNA from MPXV strains Zr-599 (Clade Ia) and TKY220091 (Clade IIb). E) Clade specific detection by the Kairo-CONAN-based lateral flow assay on the 150 aM the genomic DNA from MPXV strains Zr-599 and TKY220091 (see also Supplementary Figure 6).

To evaluate sequence-specific cleavage activity for each Clade, we generated 120-base DNA fragments and validated their Clade-specific detection capability. All crRNAs showed robust cleavage signals, confirming the assay’s accuracy, and we were able to select the crRNAs that demonstrated the strongest signals (Fig. 5B). We further proceeded to validate the Kairo-CONAN platform using genomic DNA from Mpox strains Zr-599 (Clade Ia) and TKY220091 (Clade IIb). When testing detection sensitivity using DNA fragments and CDC RT-qPCR primer sets ^25^, we achieved detection sensitivity at the 100 aM level (Fig. 5C). After quantifying the genomic DNA amounts of Zr-599 (Clade Ia) and TKY220091 using qPCR standard curves, we validated the detection specificity and sensitivity of each target-specific crRNA. The common crRNA #1 detected both RPA samples of Clade Ia and IIb MPXV at the aM level. Additionally, Clade Ia-specific crRNA showed collateral cleavage only with Zr599, while Clade Ib-specific crRNA reacted only with synthetic DNA, demonstrating detection sensitivity in the tens of aM range, comparable to qPCR sensitivity (Fig. 5D). For practical field applications, lateral flow strips enabled detection of low viral DNA concentrations at the 150 aM range (Fig. 5E and Supplementary Figs. 6A, 6B and 6C). These results confirm that Kairo-CONAN aligns with the G7’s 100DM framework and represents a valuable diagnostic tool for Mpox outbreak response in resource-limited environments.

## Discussion

The rapid and global reemergence of Mpox has underscored the pressing need for effective, field-deployable diagnostic tools that can operate reliably under outbreak conditions, particularly in regions with limited infrastructure^3,4^. This study presents Kairo-CONAN, a CRISPR-Cas3-based diagnostic platform optimized for Mpox detection in diverse environments. We also established Clade-specific detection for Mpox using crRNAs targeting Clades Ia, Ib, and IIb. While MPXV DNA samples validated detection for Clades Ia and IIb, Clade Ib could not be tested due to the unavailability of appropriate samples ^23,24^. By leveraging CRISPR-Cas3’s unique DNA-targeting properties, Kairo-CONAN achieves high sensitivity and specificity, fulfilling key diagnostic criteria in outbreak settings.

The device-free design, incorporating a disposable hand warmer (Kairo) as a sustainable heat source ^17^ and freeze-dried reagents for stability ^18^, addresses logistical challenges in low-resource settings. Lyophilized CRISPR-Cas3 components retainedhigh activity after room-temperature storage (Fig. 3F), overcoming the limitations of traditional PCR-based diagnostics that require freezing. Kairo-CONAN’s versatility was demonstrated in detecting RNA viruses, such as SARS-CoV-2, using RT-RPA (Supplementary Fig. 2A). Its compatibility with Cas12a and Cas13 further broadens its applicability to various nucleic acid targets ^12–16,26^, supporting pandemic preparedness across diverse pathogens.

In conclusion, Kairo-CONAN represents a significant advancement in portable, high-sensitivity diagnostics for Mpox, with broader applications for RNA and DNA viruses. By addressing logistical and environmental challenges, this platform aligns with the G7’s 100 Days Mission, offering a scalable and adaptable solution for outbreak response and pandemic preparedness (https://ippsecretariat.org/). Its eco-friendly design, high specificity, and field adaptability position Kairo-CONAN as a transformative tool for global health initiatives aimed at rapid disease detection and containment.

## Methods

### Protein synthesis

Cas3, mixture of Cascade components, and recombinant Cascade were synthesized as previously described ^16,21,22^. Briefly, EcoCas3 cDNA was cloned with an octa-histidine tag and a six asparagine-histidine repeat tag into a pFastbac-1 plasmid (Thermo Fisher Scientific, Waltham, Massachusetts, USA) according to the manufacturer’s instructions. The TEV protease recognition site was also inserted between the tags and EcoCas3 to enable tag removal.

For the production of mixture of Cascade components, a plasmid was designed in which each Cascade component gene was cloned into pFastbac-1 by incorporating NLS sequences at the N- and C-terminal and a HisTag sequence at either the N- or C-terminal, linked by 2A-peptide. Expression and purification of mixture of Cascade components in Sf9 was performed by Bac to bac baculovirus expression system using SF9 insect cell (Thermo Fisher Scientific Inc., Carlsbad, CA). 2A tandemed Cascade genes encoded plasmid were transformed into DH10bac competent cell. Purified each bacmid was transfected to SF9 cells, and each high titer baculoviruses were acquired by repeatedly infecting new cells with baculovirus in the culture supernatant. Acquired baculovirus added to SF9 culture cells at two multiplicity of infection (MOI), then cultured at 28°C for 24 hrs. After the infection, the culture temperature was lowered to 20°C for 3 days for protein expression. Collected Sf9 cells were collected and stored at -80°C until purification use. After ultrasonic disruption of cells, homogenized samples were ultracentrifuged and collected the supernatant including Cas proteins. Cas proteins in the supernatant were purified using nickel affinity resin (Ni-NTA, Qiagen). SuperdexTM 200 increase 10/300 GL (Cytiva Tokyo, Japan) was used for gel filtration chromatography.

The recombinant Cascade for EMX1 were purified in accordance with previously reported methods^22,27,28^. Briefly, the recombinant Cascade ribonucleoproteins (RNPs) were expressed in JM109 (DE3) by co-transformation with three plasmids: one plasmid encoding a hexahistidine tag and HRV 3C protease recognition site in the N-terminus of Cas11 (plasmid pCDFDuet-1); one plasmid containing the Cascade genes encoding Cas5, Cas6, Cas7, Cas8, and Cas11 (plasmid pRSFDuet-1); and the final plasmid encoding crRNA (pACYCDuet-1). The transformed bacteria were cultured in 2xYT medium at 37 °C with 130 rpm. After the OD_600_ became 0.6 to 0.8, we added IPTG (final concentration 0.4 mM) and cultured at 26°C with 110 rpm for 16 hours. The expressed Cascade-crRNA RNPs were purified by Ni-NTA resin. After removing the hexahistidine tag using HRV 3C protease, the recombinant Cascade RNPs were further purified by size-exclusion chromatography in 350 mM NaCl, 1 mM DTT, and 20 mM HEPES-Na (pH 7.0) and size-evaluated by SDS-PAGE.

### Preparation of DNA and RNA

For the Cascade/Cas3 activator templates, DNA fragments of hEMX1 and MPXV variants (which include a target site) were designed and purchased from Integrated DNA Technologies (IDT) or AZENTA Life Sciences (Tokyo) (Supplementary Table 1). Monkeypox virus genomic DNA was prepared as previously described^29^. Two MPXV strains, Zr-599 (Congo Basin strain, clade I) and the 2022 outbreak strain (MPXV/human/Japan/Tokyo/TKY220091/2022, clade IIb, Genbank accession no. LC722946.1), were propagated using VeroE6 cells (kindly gifted by Tokyo Metropolitan Institute of Public Health) and DNA was extracted. Cov2 RNA was prepared as previously described^16^ . Viral RNAs from SARS-CoV-2 were prepared according to the established protocol from the National Institute of Infectious Diseases in Japan ^30^. Viral RNAs were purified from an infected TMPRSS2-expressing VeroE6 cell line using the QIAamp Viral RNA Mini Kit (QIAGEN) according to the manufacturer’s protocol. The primers used for PCR, qPCR or RPA are listed in Supplementary Table 2. The probes were designed with the addition of FAM or HEX and were purchased from IDT or AZENTA (Supplementary Table 3).

### Design and preparation of crRNA

Cascade/Cas3 target sites were based on the human EMX1 gene, and the N gene from SARS-CoV-2, regions common to MPXV strains, and regions characteristic of each MPXV strain (Supplementary Table 4). crRNAs were purchased from Integrated DNA Technologies. Before CONAN reaction, each RNA and Cascade protein were mixed and incubated at 37°C for 10 minutes.

### Preparation of freeze-dried Cas3 and Cascade protein

Cas3 and/or Cascade protein was mixed in 20 µL freeze-drying buffer containing 10% (w/v) D-trehalose, 60 mM KCl, 10 mM MgCl_2_, 10 μM CoCl_2_, 5 mM HEPES-KOH (pH 7.5) and stored at −80°C before the freeze-drying process. After pre-freezing, freeze-drying was performed for 20-96 h with freeze dry system (Versatile and flexible Freeze Dryer; TOKYO RIKAKIKAI CO., LTD.).

### Recombinase Polymerase Amplification (RPA)

RPA of target regions for CONAN reaction was performed using TwistAmp Basic kit (TwistDx, Maidenhead, UK). RPA reaction solutions were prepared following the manufacturer’s instructions. Briefly, freeze-dried pellets were dissolved with a solution containing primers and Rehydration buffer, template DNA was added, and 280 mM Magnesium Acetate was added and and incubated using a thermal cycler at 37 °C or Kairo for 20-40 min.

### Nucleic acid detection using CONAN

CONAN were performed as previously described ^16^. Briefly, DNA templates containing DNA fragments, PCR products, RPA products were added to 100 nM Cascade-crRNA complex, 400 nM Cas3 and 1 mM ATP, CRISPR-Cas3 system working buffer (60 mM KCl, 10 mM MgCl_2_, 10 μM CoCl_2_, 5 mM HEPES-KOH pH 7.5). Single-stranded DNA probes were added at a final concentration of 1 µM. CONAN reaction solutions were incubated at 37 °C for 10-30 min with the thermal cycler or qPCR system. In case of using a kairo, after opening the kairo bag and shaking it 5 times, a 200 µL microtube containing CONAN reaction solution was sandwiched between the kairo and incubated for 10 min. Changes in fluorescence values associated with cleavage of single-stranded DNA probes were measured over time or at endpoints using CFX Connect (Bio-Rad Laboratories, Hercules, CA).

### Detection of MPXV by qPCR

The concentrations of MPXV genomes were quantified using a SsoAdvanced Universal SYBR Green Supermix (Bio-Rad) and CFX Connect. The cycling conditions were as follows: 94 °C for 2 min, followed by 40 cycles of 10 sec at 98 °C and 15 sec at 60 °C. Primers previously reported ^25^ were used (Supplementary Table 2).

### Detection of CONAN reactions by lateral flow assay

For lateral flow assays, DNA template was added to 50 nM Cascade-crRNA complex, 200 nM Cas3 and 1 mM ATP, CRISPR-Cas3 system working buffer (60 mM KCl, 10 mM MgCl_2_, 10 μM CoCl_2_, 5 mM HEPES-KOH pH 7.5). After 2µL of DNA template was added to a total of 17µL of CONAN reaction solution, 1µL of biotin-containing single-stranded DNA probes were added at a final concentration of 250 nM. CONAN reaction solutions were incubated with a thermal cycler which is set at 37°C or Kairo for 10 min. After adding 80 µL of the Milenia GenLine Dipstick Assay Buffer provided with the Lateral Flow Kit (Millenia GenLine HybriDetect, milenia biotec), a lateral flow strip was added to the reaction tube and the appearance of the test band was observed after 3 minutes. The test band intensities on the lateral flow strips were quantified by the gray values using the ImageJ tool.

### Quantification and statistical analysis

Statistical analyses were performed using GraphPad Prism 10 software (GraphPad Software). Image analysis was performed using the ImageJ tool. All data was reported as mean ± standard deviation as described where applicable. Statistical significance was tested using the two-way ANOVA with Sidak’s posthoc test unless otherwise noted and significance was reported as \**P* < 0.05, \*\**P* < 0.01, \*\*\**P* < 0.001, \*\*\*\**P* < 0.0001.

### Data availability

The research findings presented in this study are supported by data included in the main text and in Supplementary Information.

## Data Availability

All data produced in the present study are contained in the manuscript.

## Acknowledgments

We thank Mr. Tatsuo Serikawa for his valuable insights and contributions to the conceptualization of this study. We are also grateful to Ms. Hiromi Taniguchi and Ms. Yuko Yamauchi at the University of Tokyo for their technical assistance with in vitro assays, as well as to Ms. Sachiko Yamamoto and Mr. Shuku Saji at the RIKEN SPring-8 Center for their support in protein extraction and purification. This research was supported in part by JSPS KAKENHI from the Ministry of Education, Culture, Sports, Science and Technology of Japan (MEXT) (grant numbers 19KK0401, 22K19238, 22H02266, 23H00367) and a grant from the Japan Agency for Medical Research and Development (AMED) (grant numbers 23ck0106807, 24bm12230009, 223fa627001, JP24jf0126002, JP243fa627001h0003, JP243fa727002, 23fk0108583, JP24fk0108690).

## Author Contributions

KY and TM conceived and supervised the study, interpreted the data, and drafted the manuscript. RH conducted the primary experiments and analyzed the data with assistance from KA. KT was responsible for the preparation and purification of all CRISPR-Cas3 proteins. KS and KJI provided insights into study design and assisted in data interpretation. All authors contributed to the manuscript, reviewed it critically, and approved the final version prior to submission.

## References

1. Mitja, O., et al. Monkeypox. Lancet 401, 60–74 (2023).

2. Gessain, A., Nakoune, E. & Yazdanpanah, Y. Monkeypox. N Engl J Med 387, 1783–1793 (2022).

3. Lim, C.K., et al. Mpox diagnostics: Review of current and emerging technologies. J Med Virol 95, e28429 (2023).

4. Titanji, B.K., Hazra, A. & Zucker, J. Mpox Clinical Presentation, Diagnostic Approaches, and Treatment Strategies: A Review. JAMA (2024).

5. Olawade, D.B., et al. Strengthening Africa’s response to Mpox (monkeypox): insights from historical outbreaks and the present global spread. Science in One Health 3(2024).

6. Ilic, I., Zivanovic Macuzic, I. & Ilic, M. Global Outbreak of Human Monkeypox in 2022: Update of Epidemiology. Trop Med Infect Dis 7(2022).

7. Gostin, L.O., Jha, A.K. & Finch, A. The Mpox Global Health Emergency — A Time for Solidarity and Equity. New England Journal of Medicine 391, 1265–1267 (2024).

8. Nakhaie, M., et al. Monkeypox virus diagnosis and laboratory testing. Rev Med Virol 33, e2404 (2023).

9. Minhaj, F.S., et al. Orthopoxvirus Testing Challenges for Persons in Populations at Low Risk or Without Known Epidemiologic Link to Monkeypox - United States, 2022. MMWR Morb Mortal Wkly Rep 71, 1155–1158 (2022).

10. Davis, I., et al. Development of a specific MPXV antigen detection immunodiagnostic assay. Front Microbiol 14, 1243523 (2023).

11. Ye, L., et al. Gold-based paper for antigen detection of monkeypox virus. Analyst 148, 985–994 (2023).

12. Kaminski, M.M., Abudayyeh, O.O., Gootenberg, J.S., Zhang, F. & Collins, J.J. CRISPR-based diagnostics. Nat Biomed Eng 5, 643–656 (2021).

13. Mohammad, N., Katkam, S.S. & Wei, Q. Recent Advances in CRISPR-Based Biosensors for Point-of-Care Pathogen Detection. CRISPR J 5, 500–516 (2022).

14. Mustafa, M.I. & Makhawi, A.M. SHERLOCK and DETECTR: CRISPR-Cas Systems as Potential Rapid Diagnostic Tools for Emerging Infectious Diseases. Journal of Clinical Microbiology 59, 10.1128/jcm.00745-00720 (2021).

15. Kostyusheva, A., et al. CRISPR-Cas systems for diagnosing infectious diseases. Methods 203, 431–446 (2022).

16. Yoshimi, K., et al. CRISPR-Cas3-based diagnostics for SARS-CoV-2 and influenza virus. iScience 25, 103830 (2022).

17. Honma, M., et al. Primary cutaneous anaplastic large cell lymphoma successfully treated with local thermotherapy using pocket hand warmers. J Dermatol 35, 748–750 (2008).

18. Kongkaew, R., Uttamapinant, C. & Patchsung, M. Point-of-care CRISPR-based Diagnostics with Premixed and Freeze-dried Reagents. J Vis Exp (2024).

19. Verma, A., et al. Mpox 2024: New variant, new challenges, and the looming pandemic. Clinical Infection in Practice 24(2024).

20. Hitchman, R.B., Possee, R.D. & King, L.A. Baculovirus expression systems for recombinant protein production in insect cells. Recent Pat Biotechnol 3, 46–54 (2009).

21. Asano, K., et al. CRISPR Diagnostics for Quantification and Rapid Diagnosis of 2 Myotonic Dystrophy Type 1 Repeat Expansion Disorders. ACS Synthetic Biology, in press (2024).

22. Yoshimi, K., et al. Dynamic mechanisms of CRISPR interference by Escherichia coli CRISPR-Cas3. Nat Commun 13, 4917 (2022).

23. Vakaniaki, E.H., et al. Sustained human outbreak of a new MPXV clade I lineage in eastern Democratic Republic of the Congo. Nat Med 30, 2791–2795 (2024).

24. Kinganda-Lusamaki, E., et al. Clade I mpox virus genomic diversity in the Democratic Republic of the Congo, 2018-2024: Predominance of zoonotic transmission. Cell (2024).

25. Li, Y., Zhao, H., Wilkins, K., Hughes, C. & Damon, I.K. Real-time PCR assays for the specific detection of monkeypox virus West African and Congo Basin strain DNA. Journal of Virological Methods 169, 223–227 (2010).

26. Zhou, J., Li, Z., Seun Olajide, J. & Wang, G. CRISPR/Cas-based nucleic acid detection strategies: Trends and challenges. Heliyon 10, e26179 (2024).

27. Hochstrasser, M.L., et al. CasA mediates Cas3-catalyzed target degradation during CRISPR RNA-guided interference. Proc Natl Acad Sci U S A 111, 6618–6623 (2014).

28. Jore, M.M., et al. Structural basis for CRISPR RNA-guided DNA recognition by Cascade. Nat Struct Mol Biol 18, 529–536 (2011).

29. Watanabe, Y., et al. Virological characterization of the 2022 outbreak-causing monkeypox virus using human keratinocytes and colon organoids. Journal of Medical Virology 95, e28827 (2023).

30. Matsuyama, S., et al. Enhanced isolation of SARS-CoV-2 by TMPRSS2-expressing cells. Proceedings of the National Academy of Sciences 117, 7001–7003 (2020).

