## Supplementary material for "Sustainable and portable CRISPR-based diagnostics for high-sensitivity Mpox detection": supp_figure_and_Table

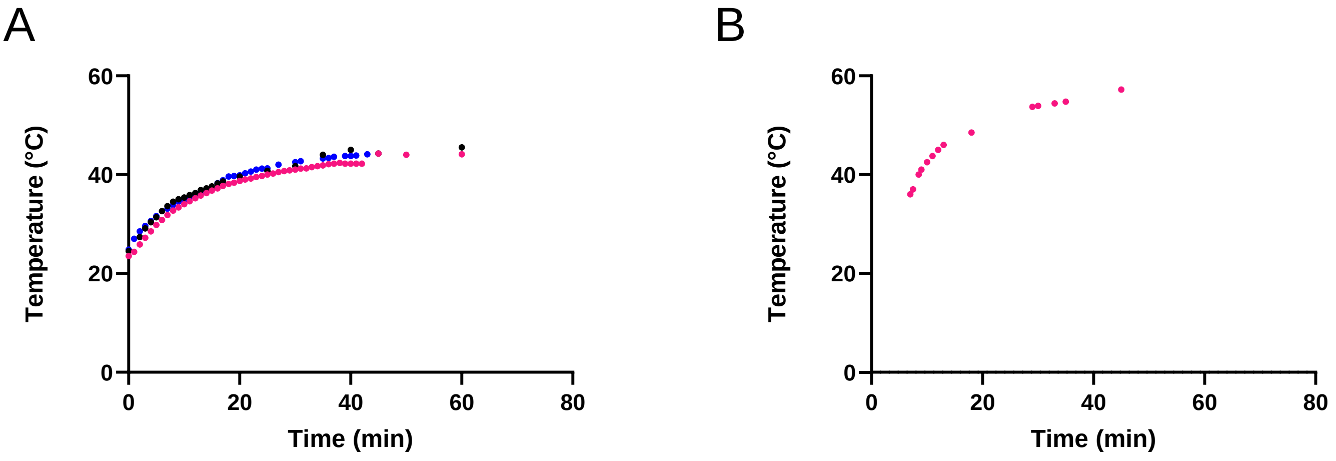


**Supplementary Figure 1. The temperature range shift of incubation using a Kairo.**

The water temperature in the microtubes sandwiched between pairs of Kairo was measured over time. Measurements were taken by opening the outer bag, holding the Kairo, and shaking it five times. (A) Incubation with Kairo model A. The three individual measurements were indicated by black, blue, and pink symbols, respectively (n = 3). (B) Incubation with Kairo model B (n = 1).

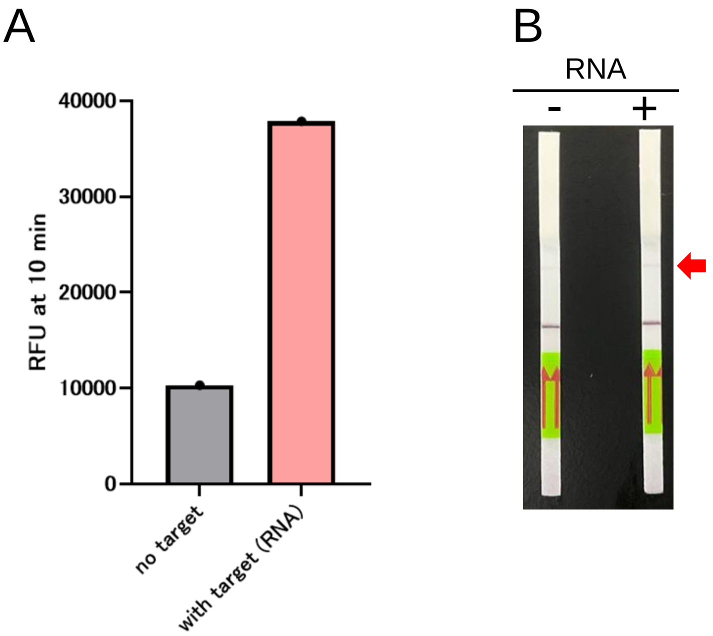


**Supplementary Figure 2. Adaptability of Kairo incubation to amplification of nucleic acids from RNA and nucleic acid detection methods with Cas12a.**

A) Collateral cleavage activity with amplified products of RT-RPA performed by Kairo incubation for 40 min with SARS-Cov2 RNA as template (n = 1). B) Lateral flow assay of DETECTR method with CRISPR-Cas12a performed by Kairo incubation against Cov2 RT-RPA products (from Supplemental figure 2A) (n = 1). Positive band position was indicated by a red arrow.

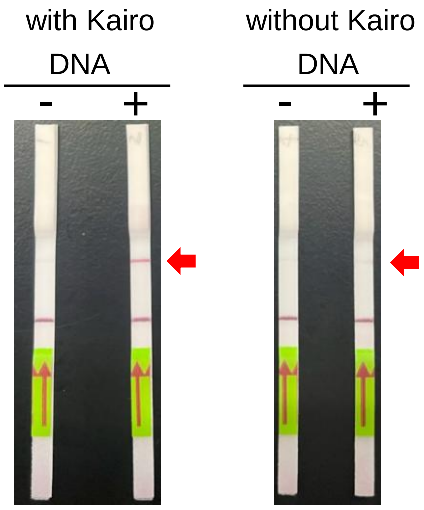


**Supplementary Figure 3. Adaptability of Kairo incubation to cold environments.**

Lateral flow detection of 500 nM EMX1 DNA fragment with (left) or without (right) Kairo incubation at 4°C (n = 1). Positive band position was indicated by a red arrow.

A

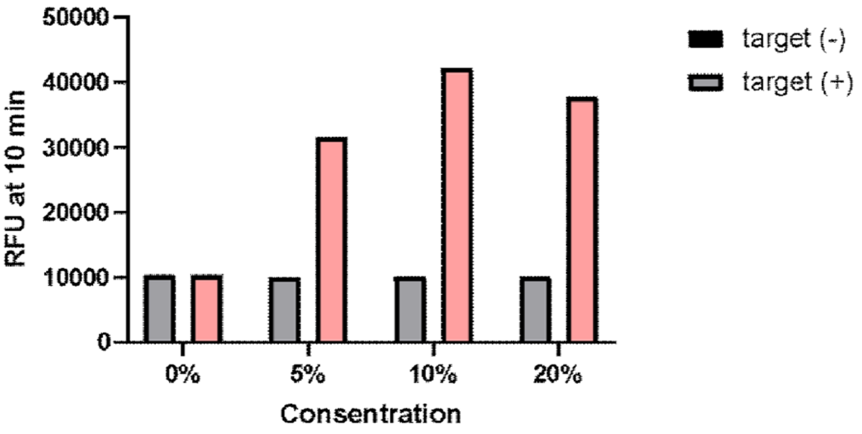


B

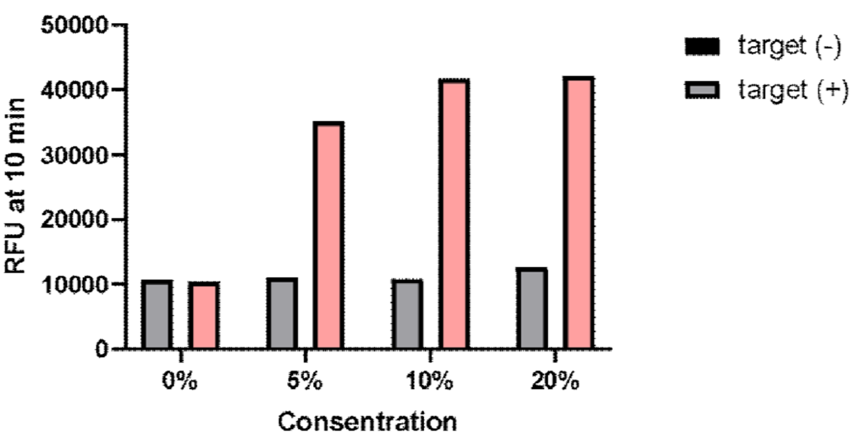


**Supplementary Figure 4. Optimization of trehalose concentration in lyophilization buffer to keep freeze-dried proteins activity**

A) Collateral cleavage activity of CRISPR-Cas3 using freeze-dried Cas3 with 0, 5, 10, 20% (w/v) trehalose against 100 nM EMX1 DNA fragment (n = 1). B) Collateral cleavage activity of CRISPR-Cas3 using freeze-dried Cascade protein with 0, 5, 10, 20% (w/v) trehalose against 100 nM EMX1 DNA fragment (n = 1).

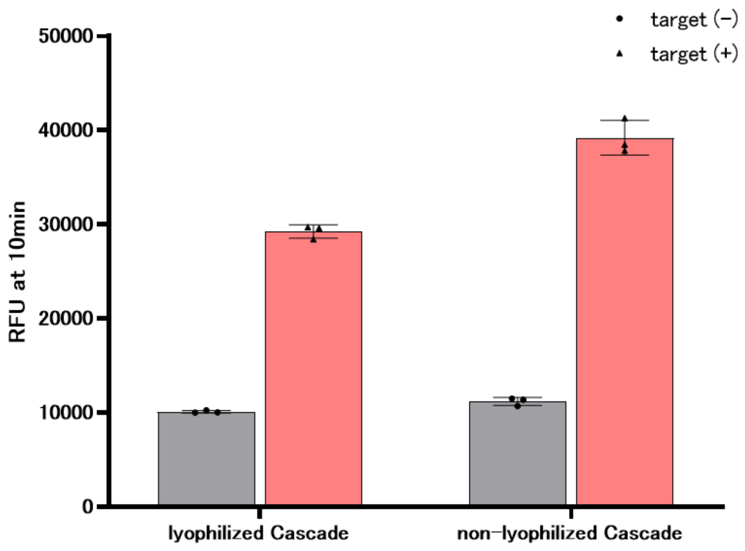


**Supplementary Figure 5. Cascade Lyophilization for MPXV Detection**

Collateral cleavage activity of CRISPR-Cas3 with the ivCascade targeting MPXV before and after lyophilization (n = 3).

Data represent means ± standard deviation.

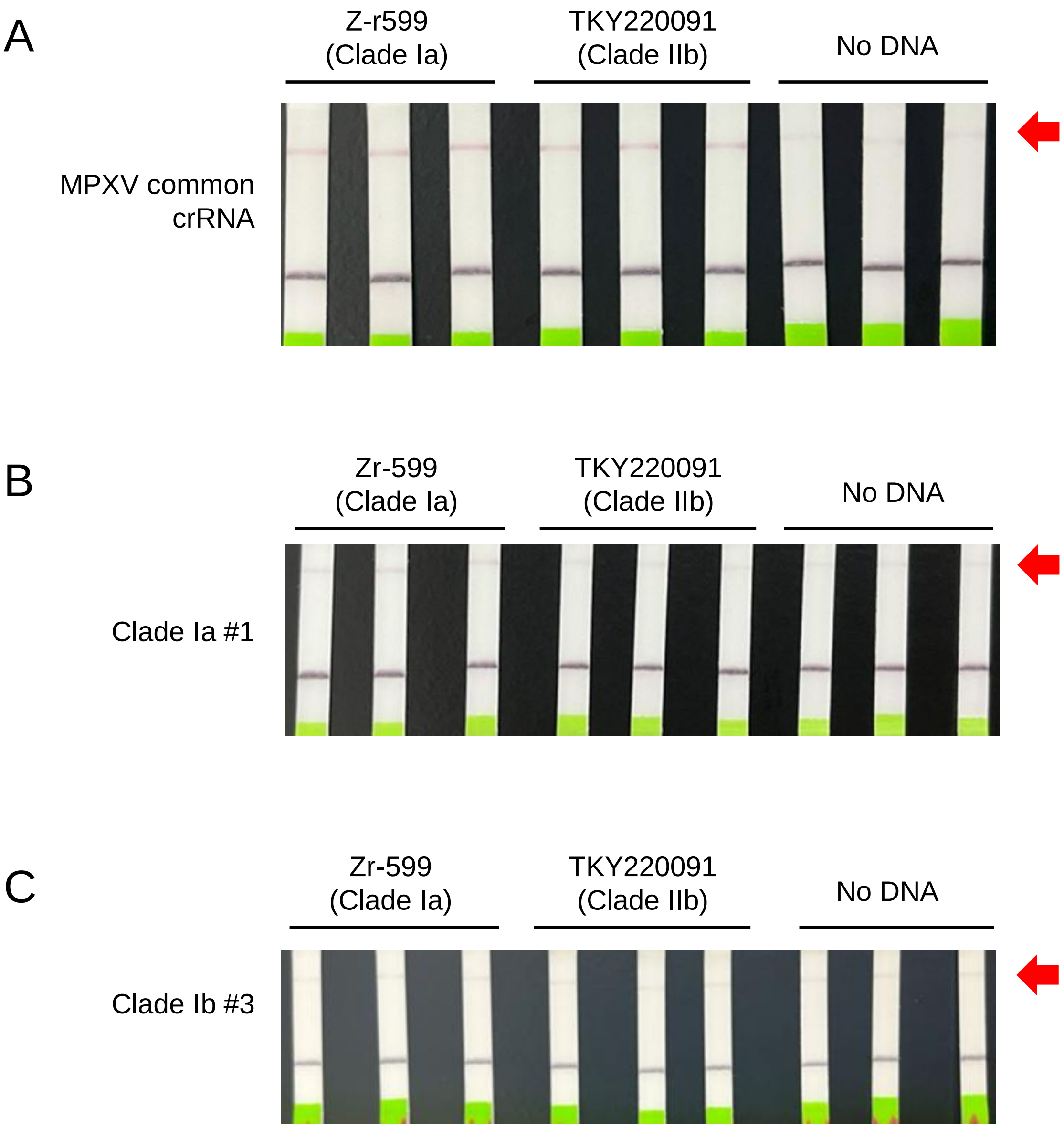


**Supplementary Figure. 6 Clade-specific detection of MPXV using Kairo and Lateral Flow Strips in the absence of electrical power.**

Lateral flow on RPA amplicon products derived from the MPXV genome (Zr-599 or TKY220091) as a template (n = 3). Incubation of RPA and CONAN were performed using Kairo. Positive band position was indicated by a red arrow. In A, RPA amplification was performed using primers targeting the common region of MPVX, while MPXV common #1 crRNA was used for CONAN. In B, RPA amplification was performed using primers targeting Clade Ia specific region, while crRNA Clade Ia #1 was used for CONAN. In C, RPA amplification was performed using primers targeting Clade Ib specific region, while crRNA Clade Ib #3 was used for CONAN.

**Supplementary Table 1. List of DNA fragment sequences**

| Name | Sequence (5' to 3') |
|---|---|
| EMX1-AAG | TGGCGCATTGCCACGAAGCAGGCCAATGGGGAGGACATCGATGTCACCTCCAATGACTAG |
| Oligo-CDC-MPXV | CCAATGGAAAATGTAAAGACAACGAATACAGAAGCCGTAATCTATGTTGTCTATCGTGTCCTCCGGGAACTTACGCTTCCAGATTATGTGATAGCAAGAC |
| MPXV-CDC fragment | TCTCTCATGTATAATAATAAACGGAAGAGATATAGCACCACATGCACCATCCAATGGAAAATGTAAAGACAACGAATACAGAAGCCGTAATCTATGTTGTCTATCGTGTCCTCCGGGAACTTACGCTTCCAGATTATGTGATAGCAAGACTAATACACAATGTACGCCGTGTGGTTCGGATACCTTTACATCTCACAATA |
| OligoDNA MPXV Ⅰb | AGCCGCTAGAAGTTTTCCGTTTGATATAGGATGTGGACATTTAACAATCTGACACGTGGGTGGATATGCGCCTGAATATCTAGGAGTGATTAAGTTTGGAAGTCTTTTCCATTTCGAAGT |

| Supplementary Table 2. List of primers | | |
|---|---|---|
| **Target** | **Name** | **Sequence (5' to 3')** |
| MPXV (qPCR) | CDC_Mpox_F | GGAAAATGTAAAGACAACGAATACAG |
| MPXV (qPCR) | CDC_Mpox_R | GCTATCACATAATCTGGAAGCGTA |
| MPXV (RPA) | MPXV-CDC_RPA_F | CTCTCATGTATAATAATAAACGGAAG |
| MPXV (RPA) | MPXV-CDC_RPA_R | GTGAGATGTAAAGGTATCCGAACCAC |
| MPXV Clade Ⅰa (RPA) | Mpox_Ⅰa_RPA_F1 | CACCAGAGTTACCAATCAACGAATATCCAC |
| MPXV Clade Ⅰa (RPA) | Mpox_Ⅰa_RPA_R1 | GAGGCACCTATTTGCGAATCTGTTAAATGCCAATC |
| MPXV Clade Ⅰb (RPA) | Mpox_Ⅰb_RPA_F1 | CCGTTTGATATAGGATGTGGACATTTAAC |
| MPXV Clade Ⅰb (RPA) | Mpox_Ⅰb_RPA_R1 | GGAAAAGACTTCCAAACTTAATCACTCC |
| SARS-CoV-2 | SARS-N2-F | TTCGGCAGACGTGGTCCAGAACAAACCCAA |
| SARS-CoV-2 | SARS-N2-R | CCTGTGTAGGTCAACCACGTTCCCGAAGGT |
| EMX1 (RPA) | EMX1_A_F2 | CACATCAACCGGTGGCGCATTGCCACG |
| EMX1 (RPA) | EMX1_A_R2 | AGCAAGCAGCACTCTGCCCTCGTGGGTTTG |

**Supplementary Table 3. List of probes used in this study**

Lowercase letters, RNA

| Name | Sequence (5' to 3') |
|---|---|
| Control probe | AGGTCGGA-ZEN-GTCAACGGATTTGGTC |
| AA reporter | uaAAgc |
| AT reporter | uaATgc |
| AG reporter | uaAGgc |
| AC reporter | uaACgc |
| TA reporter | uaTAgc |
| TT reporter | uaTTgc |
| TG reporter | uaTGgc |
| TC reporter | uaTCgc |
| GA reporter | uaGAgc |
| GT reporter | uaGTgc |
| GG reporter | uaGGgc |
| GC reporter | uaGCgc |
| CA reporter | uaCAgc |
| CT reporter | uaCTgc |
| CG reporter | uaCGgc |
| CC reporter | uaCCgc |
| pAC-6 reporter | ACACAC |
| pAC-12reporter | ACACACACACAC |
| pAC-18reporter | ACACACACACACACACAC |
| pAC-24reporter | ACACACACACACACACACACACAC |
| pA reporter | AAAAAA |
| pT reporter | TTTTTT |
| pG reporter | GGGGGG |
| pC reporter | CCCCCC |
| pAT | ATATAT |
| pAC | ACACAC |
| pAG | AGAGAG |
| pTC | TCTCTC |
| pTG | TGTGTG |
| pGC | GCGCGC |
| pCAreporter | CACACA |

**Supplementary Table 4. List of crRNA sequences**

| Name | Sequence (5' to 3') |
|---|---|
| hEMX1 | GUGUUCCCCGCGCCAGCGGGGAUAAACCGCAGGCCAAUGGGGAGGACA<br>UCGAUGUCACCUCGUGUUCCCCGCGCCAGCGGGGAUAAACCG |
| RSR-crRNA-CDC_MPXV 1 (common #1) | GUGUUCCCCGCGCCAGCGGGGAUAAACCGCCGUAAUCUAUGUUGUCUA<br>UCGUGUCCUCCGGGUGUUCCCCGCGCCAGCGGGGAUAAACCG |
| RSR-crRNA_MPXV I a #1 | GUGUUCCCCGCGCCAGCGGGGAUAAACCGUUACAAUGCUCCCAUCGAU<br>AUAAAAAUCCUCGGUGUUCCCCGCGCCAGCGGGGAUAAACCG |
| RSR-crRNA_MPXV I a #2 | GUGUUCCCCGCGCCAGCGGGGAUAAACCGACAUAACGGAUACGAGGAU<br>UUUUAUAUCGAUGGUGUUCCCCGCGCCAGCGGGGAUAAACCG |
| RSR-crRNA_MPXV I b #1 | GUGUUCCCCGCGCCAGCGGGGAUAAACCGCGCAUAUCCACCCACGUGU<br>CAGAUUGUUAAAUGUGUUCCCCGCGCCAGCGGGGAUAAACCG |
| RSR-crRNA_MPXV I b #2 | GUGUUCCCCGCGCCAGCGGGGAUAAACCGAAUCUGACACGUGGGUGGA<br>UAUGCGCCUGAAUGUGUUCCCCGCGCCAGCGGGGAUAAACCG |
| RSR-crRNA_MPXV I b #3 | GUGUUCCCCGCGCCAGCGGGGAUAAACCGAUAUUCAGGCGCAUAUCCA<br>CCCACGUGUCAGAGUGUUCCCCGCGCCAGCGGGGAUAAACCG |
